# Modelling the epidemic 2019-nCoV event in Italy: a preliminary note

**DOI:** 10.1101/2020.03.14.20034884

**Authors:** Marco Claudio Traini, Carla Caponi, Giuseppe Vittorio De Socio

## Abstract

An analysis of the time evolution of the 2019-nCoV outbreak event in Italy is proposed and is based on the preliminary data at disposal (till March 11th, 2020) on one side, and on an epidemiological model recently used to describe the same epidemic event in the Wuhan region (February 2020) on the other side. The equations of the model include the description of compartments like Susceptible (*S*), exposed (*E*), infectious but not yet symptomatic (pre-symptomatic) (*A*), infectious with symptoms (*I*), hospitalized (*H*) and recovered (*R*). Further stratification includes quarantined susceptible (*S*_*q*_), isolated exposed (*E*_*q*_) and isolated infected (*I*_*q*_) compartments. The equations are numerically solved for boundary (initial) conditions tuned on the Italian event. The rôle of quarantine is specifically emphasized and supports the strategies adopted providing a numerical description of the effects.

## I. INTRODUCTION

Late December 2019 health facilities reported cluster of patients with pneumonia of unknown origin epidemiologically linked to a seafood and wet animal wholesale market in Wuhan China. A new previous unknown betacoronavirus was discovered and identified as the etiologic agent of this new pneumonia diagnosed in Wuhan [1]. Coronaviruses (CoV) are a large family of viruses that cause illness ranging from the common cold to more severe diseases. The new virus was provisory named 2019-nCoV by World Health Organizzation on 12 January 2020 [2] and sun after the Coronavirus Study Group (CSG) based on phylogeny, taxonomy and established practice, formally recognized this virus as a sister to severe acute respiratory syndrome coronaviruses (SARS-CoVs) of the species Severe acute respiratory syndrome-related coronavirus and designates it as severe acute respiratory syndrome coronavirus 2 (SARS-CoV-2) [3]. The outbreak of the SARS-CoV-2 due to the global spread has been defined pandemia on March 11, 2020. In Europe Italy is becoming a particularly alarming and interesting place to study the evolution of the epidemic also thanks to the rapid reaction of the Italian Health organizations and the relevant control measure to prevent transmission adopted. Based on Chinese experience and the estimation of transmission rate published by Tang B *et al*. [4]. Here we develop models for the evolution of the SARS-CoV-2 during the early stages of transmission in Italy; models which may be useful for inference, forecasting or scenario analysis. Despite the fact that epidemic is changing rapidly and our results have been considered preliminary, the models we are using are considered strongly predictive and useful for the interpretation of such an unexpected event in a country like Italy.

## II. DETERMINISTIC MODELS

### A. The *SEIR* model and its limiting approximation

In the *SEIR* framework [5], individuals in the population are classified according to their infectious status: Susceptible (*S*), Exposed (*E*) (infected but not infectious), Infectious (*I*) and Recovered (*R*). In the case of a new infection (as in the present case) the population has no prior immunity, consequently the population starts out at the disease-free equilibrium (*S* ≈*N, E* = 0, *I* = 0, *R* = 0), where *N* is the total population size, and the dynamics are determined by the following equations describing the rates of change of each simplified classes:

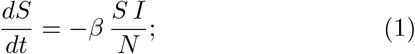

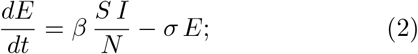

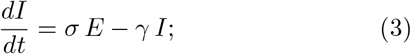

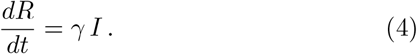

After introduction of an infectious individual (and if the basic reproductive ratio, *R*_0_ = *β/γ*, is greater than 1), the infection develops through the population by contacts between susceptible and infected individuals to sustain further transmission. Neglecting background birth and death processes (because of the short period analyzed w.r.t. the typical vital periods), the population eventually reaches the following state (*S* = *S*_∞_, *E* = 0, *I* = 0, *R* = *N* − *S*_∞_), where *S*_∞_ is defined as the solution to *R*_0_(*S*_∞_*/N* − 1) = ln(*S*_∞_*/N*) for large *N*.

The *SIR* model is a limiting case of the previous framework and assumes *E* + *I* → *I* at any time, without distinguishing the classes *E* and *I*. The previous equations reduce to

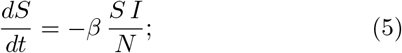

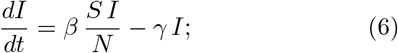

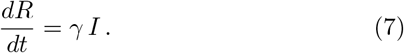

In Fig. 1 we show the available data [6, 7] for the initial period February 20th - March 11th (21 days) and their comparison with the simplest *SIR* approximation (5), (6), and (7) (iteratively solved by means of a MATLAB code) for a first intuitive analysis.

**FIG. 1.**
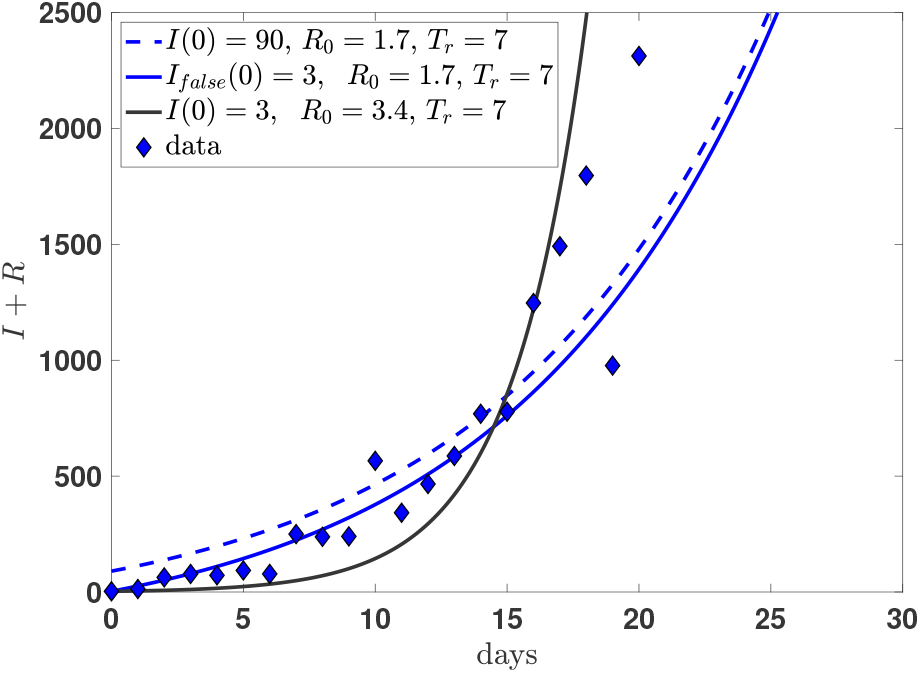
(color on line). The data cases as detected in Italy in the period February 20th (day=0) and March 11th (day = 20) (Ministero della Salute [6, 7]), compared with the prediction of few deterministic models, see text.

The continuous black curve shows the solution assuming a *I*(0) = 3 number of infectious individuals at day=0, exactly equivalent to the official detected cases at the same day. No attention is paid to a best fit of the data, also because they are rather sparse, however the quality of the simple (*SIR*) model is evident also at the first stages of the epidemic. The parameter values sound also reasonable: with a recovering average time *T*_*r*_ = 7 days (*γ* = 1*/T*_*r*_) and *R*_0_ = 3.4 and no further corrections: a brilliant example of the predictive force of the *SIR* model. However Fig. 1 has a more specific aim: showing the rather delicate rôle played by a possible underestimation of the infectious individual at the early stages of the infection outbreak. In fact one can assumes a different (possible) scenario based on the difficulty of a precise estimation of the initial number of infectious individuals. Such a possible scenario is illustrated by the the full blue curve showing a prediction which results from a misleading estimation of *I*(0). The effective number of initial infectious is 30 times larger (*I*(0) = 90), but the curve has been translated to a fictitious *Ī*(0) = 3, the reveled number, from the dashed mathematical result. A change in *R*_0_ from *R*_0_ = 3.4 to *R*_0_ = 1.7 would give a reasonable fit of the early stage of the outbreak. The effect is rather well known as discussed in ref. [8] in the case of the epidemic event in China. In that case a proposed exponential fit of the precocious data (January 10th to January 24th) results in a values of *R*_0_ ≈3.58 which is reduced to *R*_0_ ≈2.24 is the associated initial condition on *I*(0) is multiplied by an increasing factor from 2 to 8. In Fig. 2 we show the time behavior of the infectious population within the simple *SIR* model of Eqs. (5)-(7) (*T*_*r*_ = 1*/γ* = 7 days, *R*_0_ = 3.4) both in logarithmic (upper panel) and linear scale (lower panel). The data are the data of Fig. 1.

**FIG. 2.**
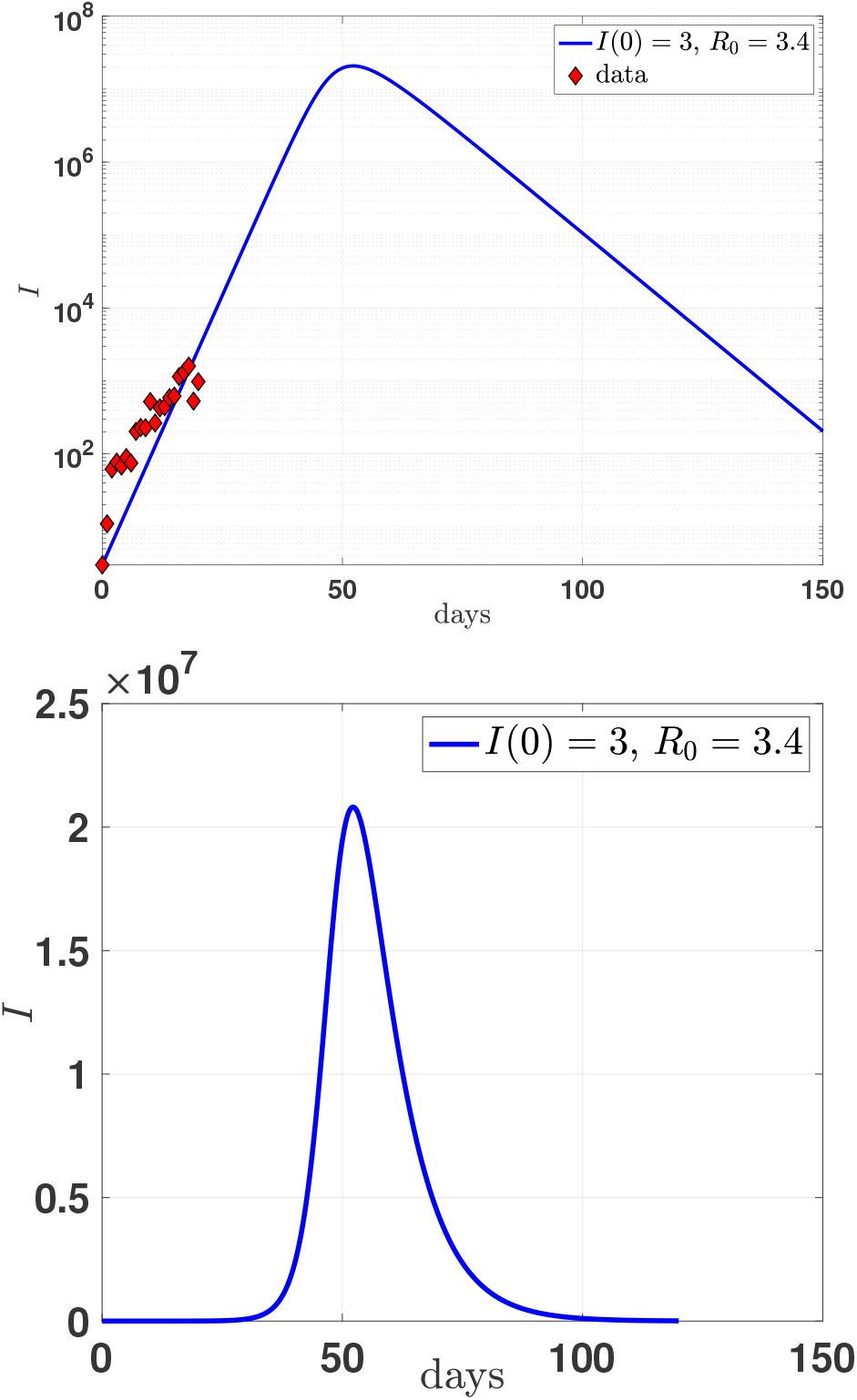
(color on line) **upper panel** Behavior (in logarithmic scale) of the number of infectious individuals in Italy within the *SIR* model and the comparison with available data till March 11th, 2020. **lower panel**: As in the upper panel but in linear scale to better appreciate the global behavior and the effective time to recover.

The exponential behavior predicted by the model in the growing part of the outbreak description is evident and can be compared with the recent fit to the data by

Bucci and Marinari in ref. [9]. While the present simplified *SIR* model predict a doubling infectious population each (roughly) 2 days, the Roma fit [9] gives an estimation of 2.6 days (see Fig. 3).

**FIG. 3.**
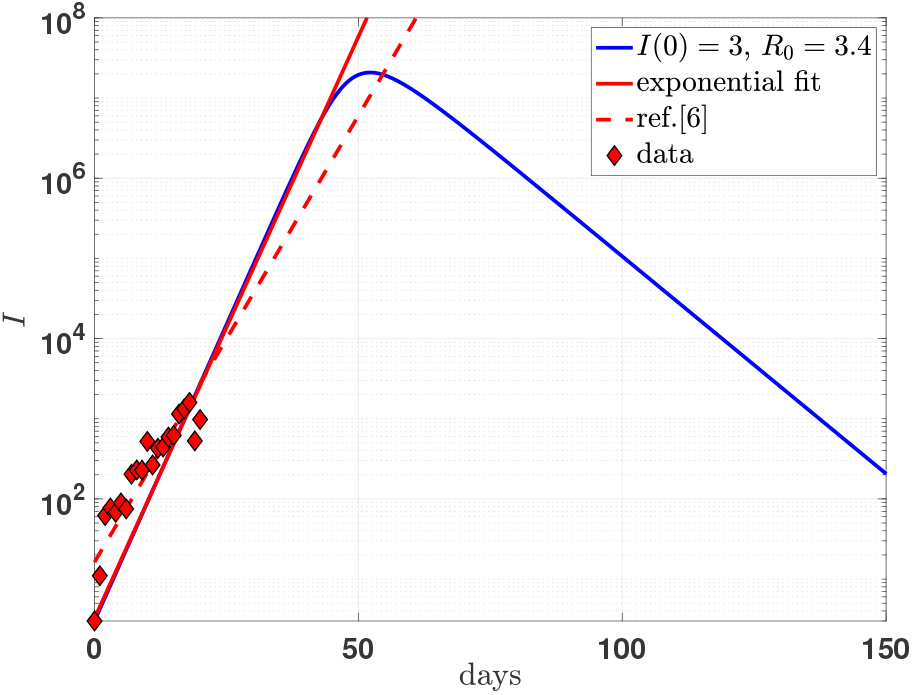
(color on line). The model predictions of Fig. 2 are compared with an exponential fit (full red line) *I* ∼ exp (*t*_*days*_*/*2.994) and compared with the fit proposed in ref.[9] (dashed red line).

### B. A Quarantined model with isolation

The model [4] is a generalized *SEIR*-type epidemiological model which incorporates appropriate compartments relevant to intervention such as quarantine, isolation and treatment. The population is stratified in Susceptible (*S*), exposed (*E*), infectious but not yet symptomatic (pre-symptomatic) (*A*), infectious with symptoms (*I*), hospitalized (*H*) and recovered (*R*). Further stratification includes quarantined susceptible (*S*_*q*_), isolated exposed (*E*_*q*_) and isolated infected (*I*_*q*_) compartments. The equations of the model are explicitly shown in Eqs. (8)-(15).

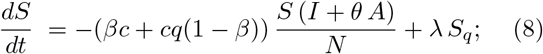

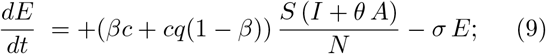

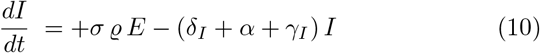

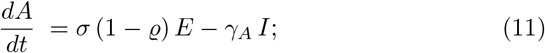

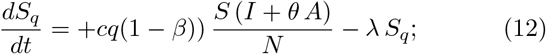

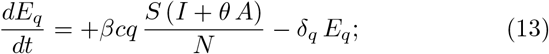

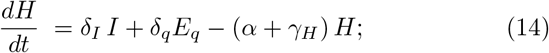

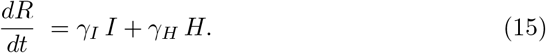

The authors of ref. [4] have calibrated the model on the data of the 2019-nCoV as emerged in Wuhan the last two months, therefore in a situations that has the same basic parameters. In table I the values of the model parameters as tuned by Tang *et al*. on the outbreak event at Wuhan are summarized and the initial conditions imposed to the solutions of the Eqs. (8)-(15) are summarized in table II for the populations localized in Wuhan and in Italy.

**TABLE I.**
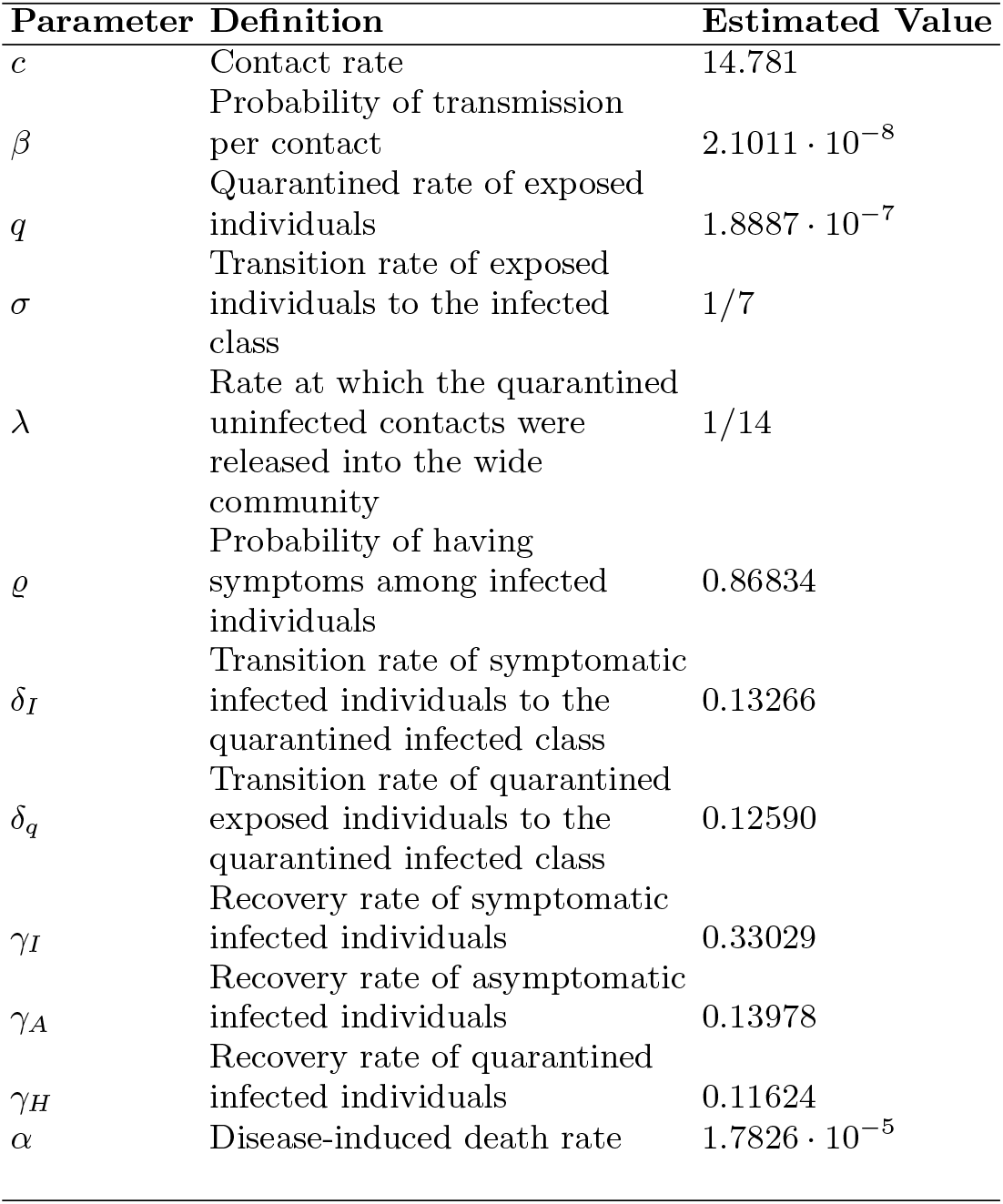
Parameters of the SEIR model description of the Wuhan outbreak. The contact rate Probability transmission has been reduced by a factor 0.14 to describe the data of the Italian event. In addition the value *θ* = 1 in Eqs. (8)-(15) has been assumed to be to reduce the model dependence.

**TABLE II.** default

| Initial values | Italy | Wuhan |
| --- | --- | --- |
| $N$ | $6 \cdot 10^7$ | $1.1 \cdot 10^7$ |
| $E(0)$ | 100 | 105 |
| $I(0)$ | 3 | 28 |
| $A(0)$ | 100 | 53 |
| $S_q(0)$ | 0 | 739 |
| $E_q(0)$ | 0 | 1 |
| $H(0)$ | 0 | 1 |
| $R(0)$ | 0 | 2 |

The results are shown in Figs 4 and 5. The drastic reduction of the infectious population is evident as well as the long tail of the distribution.

**FIG. 4.**
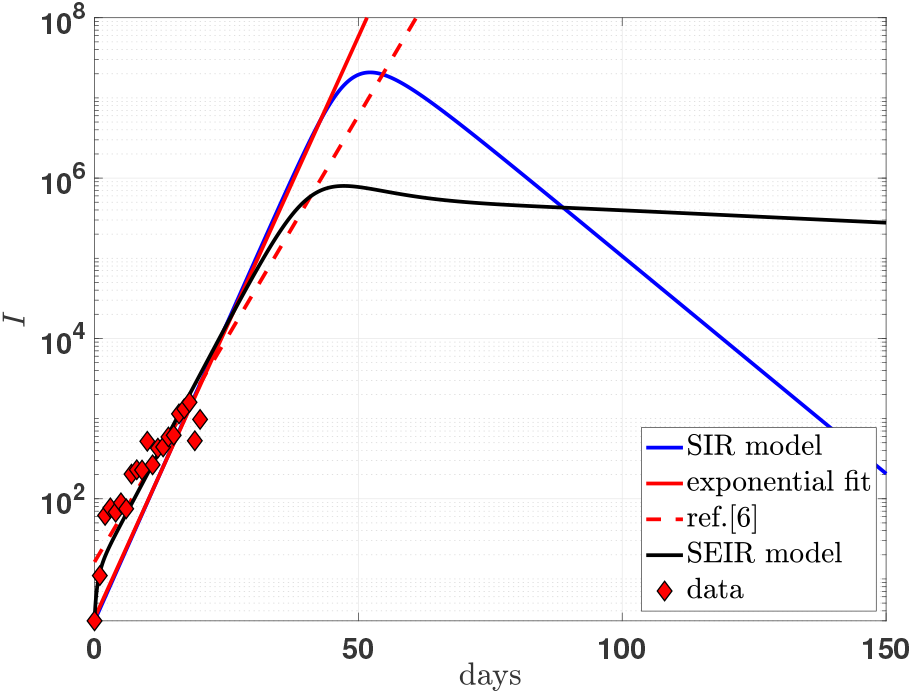
(color on line). Number of infectious individuals for the 2019-nCoV outbreak in Italy. The SEIR model predictions (full black line) are compared with the simplified results of the SIR model of Fig. 3 as well as the proposed exponential fit of Bucci and Marinari [9] (see Figure caption). The maximum values are drastically reduced, at the cost of a long tail in the distribution.

**FIG. 5.**
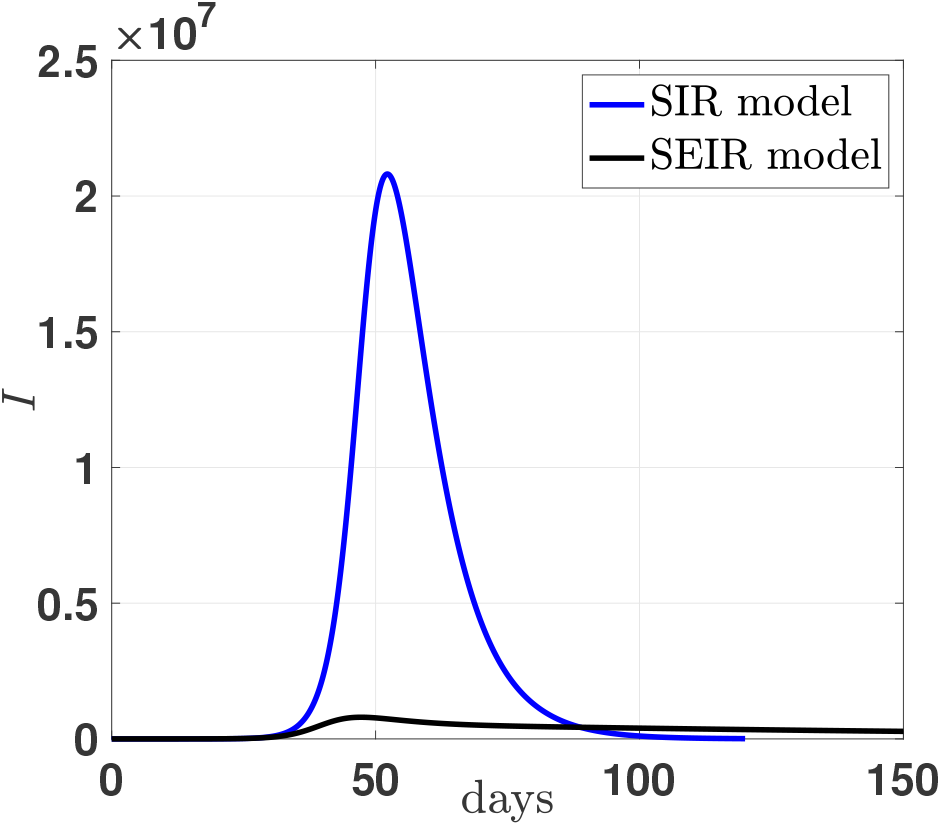
(color on line). The time evolution of the infectious population in the Italian outbreak. The linear scale emphasize the drastic reduction induced by isolation, quarantine and intense tracing as described by the recent formulation of a SEIR model based on the Wuhan database (see ref. [4]).

The quarantine imposed to the infectious individuals, the effects of an intensive contact tracing and isolation reduce the maximum value of the distribution by a factor ∼ 26 (see Fig. 5).

## III. CONCLUSIONS

The present calculation strongly supports the interventions and restrictions adopted in Italy to reduce the outbreak of the infectious population of the 2019-nCoV.

## Data Availability

data from the national site of Health Minister http://www.salute.gov.it/nuovocoronavirus

## Appendix A: Parameters and tables

We summarize in the table I of the present appendix the parameters of the SEIR-type model as proposed by Tang *et al*. in ref. [4]. Table II is devoted to summarize the initial conditions imposed to the SEIR solutions in the numerical calculation.

## Notes

### Competing Interest Statement

The authors have declared no competing interest.

### Funding Statement

No Funding received for the present research

